# Presence of cells in the polyaneuploid cancer cell (PACC) state predicts risk of recurrence in prostate cancer

**DOI:** 10.1101/2022.09.19.22280051

**Authors:** Levent Trabzonlu, Kenneth J. Pienta, Bruce J. Trock, Angelo M. De Marzo, Sarah R. Amend

**Affiliations:** Cancer Ecology Center, The Brady Urological Institute, Johns Hopkins University School of Medicine; The Brady Urological Institute, Johns Hopkins School of Medicine, Baltimore, MD 21287; Department of Pathology and Laboratory Medicine, Loyola University Medical Center, Maywood, IL 60153; Departments of Pathology, Urology and Oncology, The Johns Hopkins University School of Medicine. The Sidney Kimmel Comprehensive Cancer Center at Johns Hopkins

## Abstract

**Background:** The non-proliferating polyaneuploid cancer cell (PACC) state is associated with therapeutic resistance in cancer. A subset of cancer cells enters the PACC state by polyploidization and acts as cancer stem cells by undergoing depolyploidization and repopulating the tumor cell population after the therapeutic stress is relieved. Our aim was to systematically assess the presence and importance of this entity in men who underwent radical prostatectomy with curative intent to treat their presumed localized PCa.

**Materials and Methods:** Men with NCCN intermediate- or high-risk PCa who underwent radical prostatectomy l from 2007-2015 and who did not receive neoadjuvant treatment were included. From the cohort of 2159 patients, the analysis focused on a subcohort of 209 patients was selected, and 38 cases. Prostate tissue microarrays (TMAs) were prepared from formalin-fixed paraffin-embedded blocks of the radical prostatectomy specimens. A total of 2807 tissue samples of matched normal/benign and cancer were arrayed in 9 TMA blocks. The presence of PACCs and the number of PACCs on each core were noted.

**Results:** The total number of PACCs and the total number of cores with PACCs were significantly correlated with increasing Gleason score (p=0.0004) and increasing CAPRA-S (p=0.004), but no other variables. In univariate proportional hazards models of metastasis-free survival, year of surgery, Gleason score (9-10 vs. 7-8), pathology stage, CAPRA-S, total PACCs, and cores positive for PACCs were all statistically significant. The multivariable models with PACCs that gave the best fit included CAPRA-S. Adding either total PACCs or cores positive for PACCs to CAPRA-S both significantly improved model fit compared to CAPRA-S alone.

**Conclusion:** Our findings show that the number of PACCs and the number of cores positive for PACCs are statistically significant prognostic factors for metastasis-free survival, after adjusting for CAPRA-S, in a case-cohort of intermediate- or high-risk men who underwent radical prostatectomy. In addition, despite the small number of men with complete data to evaluate time to mCRPC, the total number of PACCs was a statistically significant predictor of mCRPC in univariate analysis, and suggested a prognostic effect even after adjusting for CAPRA-S.

## Introduction

Therapeutic resistance in cancer is generally attributed to the existence of resistant cell clones ^1–3^. The resistant clones are thought to be generated through either intrinsic genetic instability resulting in tumor cell heterogeneity (TCH) or a cancer stem cell (CSC) population ^4–7^. The resistant clones allow for regrowth of the cancer cell population after treatment insult. There is a growing body of evidence that the phenomenon of therapeutic resistance may be explained by a poorly recognized but distinct cell state. This cell state is documented as the non-proliferating polyaneuploid cancer cell (PACC) state, induced by tumor microenvironmental (intrinsic) or therapeutic (extrinsic) stress ^8–10^. A doubling of a cancer cell’s aneuploid genome combined with an exit from the cell cycle enables the PACC state. The cell state can exist for an extended period of time. In response to stress, a subset of cancer cells enter the PACC state by accessing an evolutionary or developmental polyploidization program. Once that stress is relieved, cells in the PACC state can act as CSCs by undergoing depolyploidization and repopulating the tumor cell population. The role of the PACC state for therapeutic resistance adds to those of CSC and TCH. Identifying cells in the PACC state in patients may have important diagnostic, prognostic, and therapeutic implications.

Large pleomorphic cancer cells with irregular nuclei have been documented in histopathologic specimens of multiple tumor types since the 1800s ^11^. These cells have been reported utilizing a variety of names: polyaneuploid cancer cells (PACCs), polyploid giant cancer cells (PGCCs), giant cancer cells (GCCs), multinucleated giant cancer cells (MNGCCs), blastomere-like cancer cells, osteoclast-like cancer cells, cells in an embryonic diapause, pleomorphic giant cells, large stem cells, and persister cells ^12–23^. Recent data indicates that these cells do not represent a different type of cancer cell within the heterogeneous tumor cell population but rather a specific non-proliferative, polyaneuploid cell state ^8–10^. In response to therapeutic stress, this cell state enables survival and is responsible for driving therapeutic resistance to nearly all available treatment regimens.

The presence of cells in the PACC state has been documented in virtually all cancer types including adenocarcinomas, transitional cell tumors, squamous cell carcinomas, leukemias, lymphomas, glioblastomas, and sarcomas ^12,24–29^. Most often observed in metastatic cancers or after treatment, their importance for patient prognosis has been understudied ^30,31^. In glioma, Qu and colleagues analyzed 76 patients and reported that the number of PACCs increased with the grade of tumors ^32^. In a study of 47 patients with anorectal melanoma, the number of PACCs was demonstrated to increase with tumor size ^33^. In laryngeal cancer, Liu and colleagues analyzed the presence of PGCCs in 102 patients and found that patients with high expression of PGCCs had a poorer prognosis ^34^. In breast cancer, Fei and colleagues analyzed 167 histopathologic specimens including benign tissue, primary breast tumors, and lymph node metastases and found the highest number of PGCCs in the lymph node metastases of the breast cancer patients ^30^. In a study of 30 patients, Gerashchenko and colleagues reported that breast tumors with higher proportion of polyploid cells was a marker of poor response to neoadjuvant chemotherapy ^35^. Lv and colleagues investigated the presence of PGCCs with budding in 80 patients with serous ovarian tumors and found that the presence of PGCCs in the primary tumor correlated with metastasis ^36^. Zhang and colleagues examined tissue from 159 patients with colorectal cancer and demonstrated that the presence of PGCCs with budding increased as tumors became more de-differentiated ^37^.

The presence of cancer cells in the PACC state has been reported in prostate cancer (PCa) ^12,38^. In an autopsy study of PCa patients who had failed multiple lines of therapy, Mannan and colleagues reported the presence of multiple cells with highly irregular polylobulated nuclei or multiple pleomorphic nuclei ^39^. Alharbi and colleagues reported a series of 30 patients with a rare variant of PCa with focal pleomorphic giant cell features that were extremely aggressive and associated with poor outcomes ^40^. This study was undertaken to systematically assess the presence and importance of cells in the PACC state in men who underwent radical prostatectomy with curative intent to treat their presumed localized PCa.

## Materials and Methods

### Patients

Men with NCCN intermediate- or high-risk PCa who underwent radical prostatectomy at Johns Hopkins Hospital from 2007-2015 and who did not receive neoadjuvant treatment were identified from the Institutional Review Board-approved Brady Urological Institute Radical Prostatectomy database. Intermediate risk was defined as clinical stage T2b-T2c, or biopsy Gleason grade groups 2-3, or PSA 10-20 ng/ml, and high risk was defined as biopsy Gleason grade groups 4-5 or clinical stage T3, or PSA>20 ng/ml (2). There were 3685 men with NCCN intermediate- or high-risk PCa with radical prostatectomy from 2007-2015 who did not receive neoadjuvant treatment. Of those, 2159 (59%) had complete follow-up for metastasis through 2015 and represented the pool from which the case-cohort sample was drawn.

### Case-cohort

The case-cohort was originally assembled to include as “cases” men with metastasis or with biochemical recurrence (BCR) with a rapid PSA doubling time (<10 months) who were also considered to be at high risk of metastasis^41^. From the cohort of 2159 patients, a subcohort of 244 patients was selected, and 115 cases (73 with BCR and rapid PSA doubling time, and 42 with metastasis). Tissue samples from these men were analyzed for PACC; samples from 307 men were informative for PACC. To focus specifically on risk of metastasis as the outcome of interest, we excluded 65 “men with BCR and rapid PSA doubling time but without metastasis who were included as “cases” in the original case-cohort. This resulting in a subcohort of 209 patients (including 8 metastasis cases), and 30 metastasis cases not in the subcohort, all of whom were informative for PACCs.

### Tissue Microarrays (TMAs)

Prostate TMAs were prepared from formalin-fixed paraffin-embedded blocks of the radical prostatectomy specimens. A total of 2807 tissue samples of matched normal/benign and cancer were arrayed in 9 TMA blocks. These TMAs were all constructed as described ^41–44^ from the index tumor (highest grade) with a 3–4 fold sampling redundancy. On H&E stained tissue sections and TMA sections, identifying PACCs is difficult since they often have indistinct cell membranes therefore making it difficult to distinguish “pseudo” multinucleation from the real ones. Also, unlike many other types of adenocarcinoma and poorly differentiated carcinomas (e.g. high grade urothelial carcinomas, non-small cell lung carcinomas), easily recognizable multinucleated or bizarrely enlarged nuclei are not readily apparent in the vast majority of even very high grade cases. Therefore, we used immunohistochemistry against EpCAM (mouse monoclonal antibody; ABCAM ab7504, Cambridge, UK) on these TMAs to visualize the epithelial cell membranes.. All TMA slides were scanned on a Hamamatsu Nanozoomer and imported into Concentriq (from Proscia). The whole slide scan files were evaluated by two pathologists (LT and AMD). PACCs were defined as the large multi-nucleated or poly-lobated cells that are at least three times the size of a neighboring tumor cell as assessed by visual inspection ^39^. The presence of PACCs and the number of PACCs on each core were noted.

### Statistical Analyses

Descriptive statistics were used to compare cases and subcohort, including Wilcoxon rank sum test for continuous variables and Fisher’s exact test or chi-square test for categorical variables. Correlations between PACCs and clinical variables were performed with linear regression or analysis of variance (ANOVA). The primary outcome was metastasis confirmed by imaging, and metastasis-free survival (MFS) was measured from the date of radical prostatectomy. Multivariable Cox proportional hazards regression models, with weights for the case-cohort design and robust variance estimator defined by Barlow (29) were fit to metastasis-free survival (MFS) to evaluate the hazard ratio (HR) and 95% confidence interval (CI) associated with PACCs, adjusted for established prognostic factors or Cancer of the Prostate Risk Assessment Post-surgical (CAPRA-S) score. The CAPRA-S score combines pathologic Gleason score, pathologic stage, surgical margin status and preoperative prostate specific antigen (PSA) in an algorithm with values ranging from 0-12; scores of 6 or higher are considered to indicate high risk of biochemical recurrence (12). Improvement in multivariable model fit for addition of PACCs was assessed with the pseudo-likelihood ratio test for the change in deviance from the full versus the reduced model (30). In addition to the primary outcome of MFS, follow-up for development of mCRPC was available for 33 of the men with metastases. Time from diagnosis of metastasis to mCRPC was analyzed using standard proportional hazards regression, and improvement in model fit was assessed with the likelihood ratio test. All analyses were performed with SAS v. 9.4 (SAS Institute, Cary, NC).

## Results

Since the great majority of prostatic adenocarcinomas do not frequently show bizarre nuclear atypia with extremely large nuclei or multinucleation that is readily apparent by H&E staining, we performed IHC against EpCAM to facilitate the recognition of cellular plasma membranes. This greatly aided in the ability to confidently identify PACCs (**Fig. 1** and **Supplemental Fig. 1**), which in this study were limited to cells with multiple nuclei bounded by a single plasma membrane. **Table 1** compares the metastasis cases to men without metastasis. Cases had the expected higher risk profile, differing significantly for all variables except age. Although many patient tumor samples had none, PACCs were significantly more frequent in cases with metastasis, 20 of 38 cases (52.6%), vs. 68 of 201 (33.8%) controls, p=0.029.

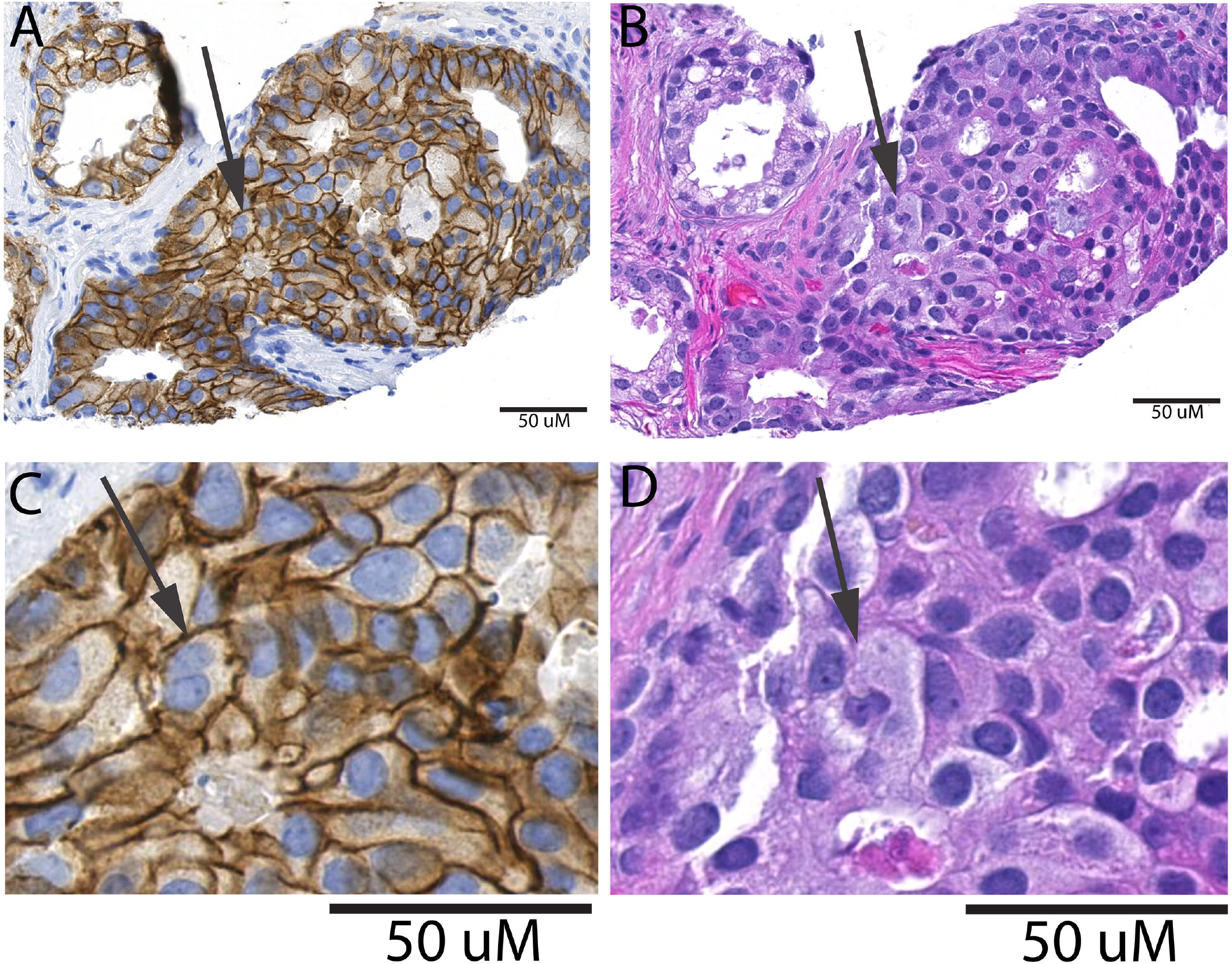

**Table 1:**
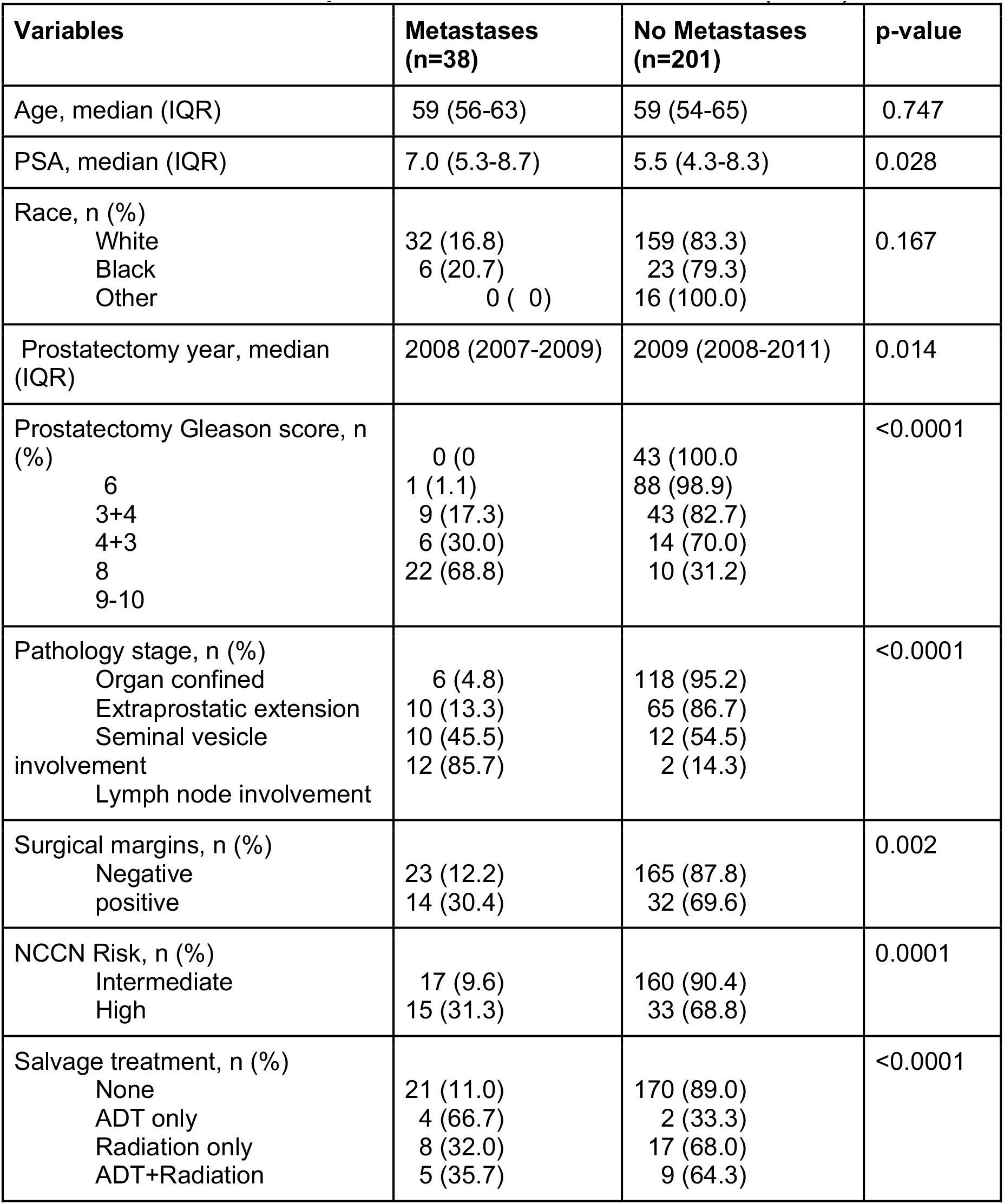
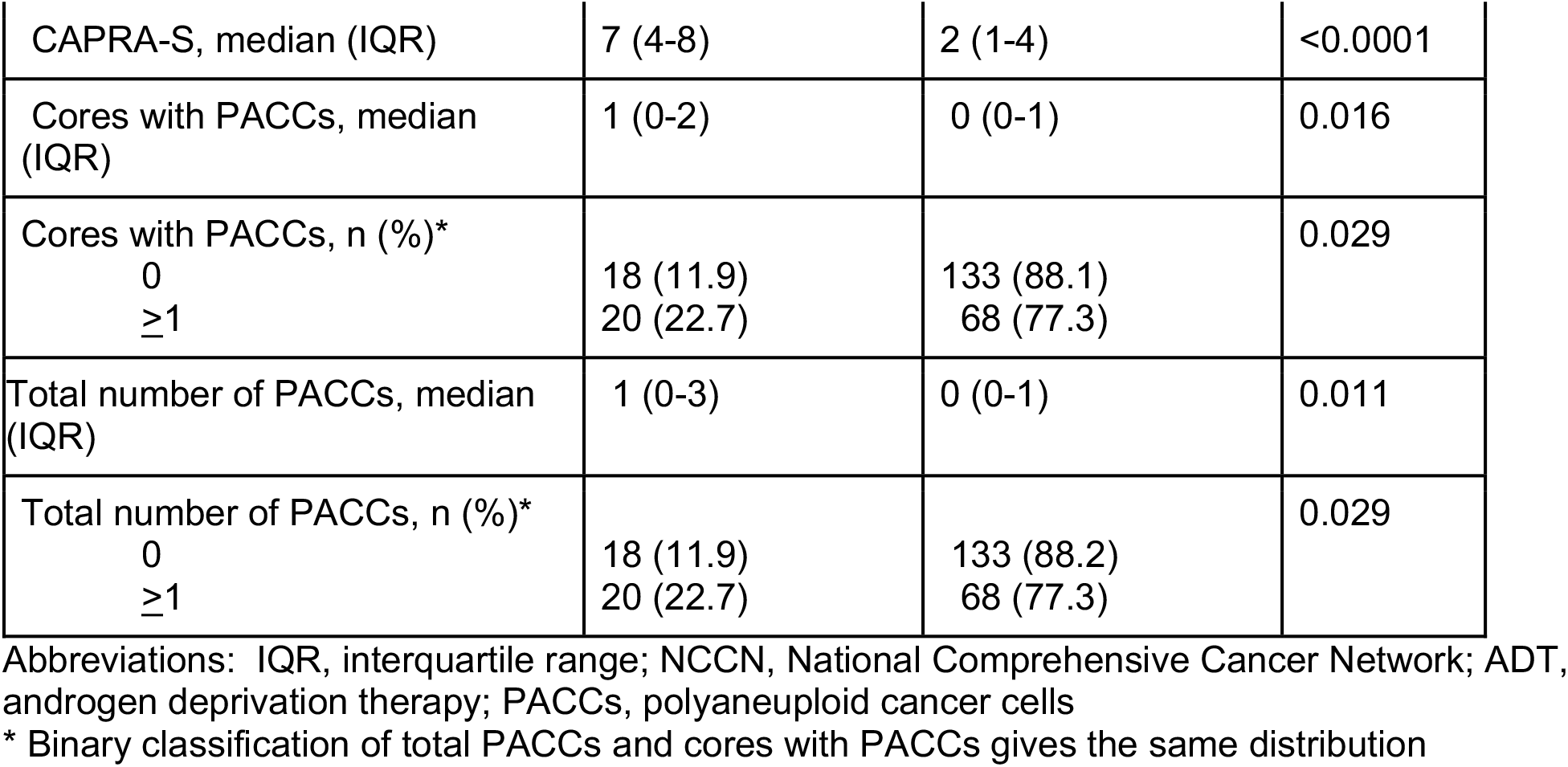
Characteristics of patients with vs. without metastases (n=239)

The total number of PACCs and the total number of cores with PACCs were significantly correlated with increasing Gleason score (p=0.0004) and increasing CAPRA-S (p=0.004), but no other variables.

In univariate proportional hazards models of MFS, year of surgery, Gleason score (9-10 vs. 7-8), pathology stage, CAPRA-S, total PACCs, and cores positive for PACCs (both evaluated as a continuous variable or dichotomized at ≥1 vs. 0) were all statistically significant (Table 2). Note that dichotomizing total PACCs and cores positive for PACCs gave the same distribution so the hazard ratios (HRs), 95% confidence intervals (CIs), and p-values are the same.

**Table 2.** Univariate proportional hazards models of metastasis-free survival in a case-cohort of intermediate and high risk men.

| Variables | HR (95% CI) | p-value |
| --- | --- | --- |
| Year of surgery | 1.50 (1.12, 2.02) | 0.007 |
| Age | 0.99 (0.93, 1.05) | 0.671 |
| PSA (per 1 ng/ml) | 1.03 (0.996, 1.07) | 0.079 |
| Gleason score<br>7-8<br>9-10 | 1.0<br>13.48 (5.40, 33.65) | <0.0001 |
| Pathologic stage<br>Organ-confined<br>Extra-prostatic extension<br>Seminal vesicle<br>involvement<br>Lymph node involvement | 1.0<br>3.95 (1.27, 12.32)<br>17.05 (3.83, 75.83)<br>61.46 (11.58, 326.33) | <0.0001 |
| CAPRA-S (per 1 unit) | 2.32 (1.79, 3.01) | <0.0001 |
| Total PACCs (per 1 PACC) | 1.38 (1.09, 1.73) | 0.006 |
| Total PACCs (≥1 vs. 0)* | 2.70 (1.24, 5.86) | 0.012 |
| Cores with PACCs (per 1 core) | 1.40 (1.004, 1.94) | 0.047 |
| Cores with PACCs (≥1 vs. 0)* | 2.70 (1.24, 5.86) | 0.012 |

Abbreviations: HR, hazard ratio; CI, confidence interval
\* Binary classification of total PACCs and cores with PACCs gives the same distribution, so results from proportional hazards model are also the same.

The multivariable models with PACCs that gave the best fit included CAPRA-S. Adding either total PACCs or cores positive for PACCs (both expressed as a continuous variable) to CAPRA-S both significantly improved model fit compared to CAPRA-S alone, based on the pseudo-likelihood ratio test (Table 3). The best-fitting model included total PACCs, HR=2.00 (95% CI 1.40, 2.87), and CAPRA-S, HR=2.51 (95% CI 1.81, 3.48).

**Table 3.** Multivariable proportional hazards models of metastasis-free survival in a case-cohort of intermediate and high risk men.

| Variables | HR (95% CI) | p-value | p-value for increase in PLRT* compared to CAPRA-S alone |
| --- | --- | --- | --- |
| Model 1 |  |  |  |
| CAPRA-S (per 1 unit) | 2.50 (1.81, 3.46) | <0.0001 | <0.0001 |
| Total PACCs (per 1 PACC) | 2.00 (1.40, 2.85) | 0.0001 |  |
| Model 2 |  |  |  |
| CAPRA-S (per 1 unit) | 2.23 (1.73, 2.88) | <0.0001 | 0.076 |
| Total PACCs (≥1 vs. 0)** | 1.97 (0.54, 7.24) | 0.306 |  |
| Model 3 |  |  |  |
| CAPRA-S (per 1 unit) | 2.35 (1.79, 3.09) | <0.0001 | 0.0003 |
| Cores with PACCs (per 1 core) | 2.15 (1.23, 3.78) | 0.007 |  |
| Model 4 |  |  |  |
| CAPRA-S (per 1 unit) | 2.23 (1.73, 2.88) | <0.0001 | 0.076 |
| Cores with PACCs (≥1 vs. 0)** | 1.97 (0.54, 7.24) | 0.306 |  |

Abbreviations: HR, hazard ratio; CI, confidence interval; PLRT, pseudo-likelihood ratio test
\* The pseudo-likelihood ratio test is based on the change in deviance when one of the PACC variables is added to a model of CAPRA-S alone.
\*\* Binary classification of total PACCs and cores with PACCs gives the same distribution, so results from proportional hazards model are also the same.

Among 38 patients with metastases, 32 had complete data on PGCCs, CAPRA-S, and time to mCRPC. Table 4 shows univariate analyses for CAPRA-S and total PACCs (expressed as a continuous variable), and the multivariable model of total PACCs and CAPRA-S. Cores positive for PACCs was not statistically significant in a univariate model or adjusted for CAPRA-S (data not shown). Total PACCs was statistically significant in the univariate model, HR=1.23 (95% CI 1.06, 1.43), p=0.007, but was no longer significant when adjusted for CAPRA-S, HR=1.17 (95% CI 0.995, 1.38), p=0.057. The small sample size resulted in a lack of statistical power, which may have influenced the result.

**Table 4.** Univariate and multivariable proportional hazards models of time from metastasis to mCRPC in a cohort of intermediate and high risk men with metastasis (n=32)

| Variables | HR (95% CI) | p-value | p-value for increase in LRT* compared to CAPRA-S alone |
| --- | --- | --- | --- |
| **Univariate** |  |  |  |
| CAPRA-S (per 1 unit) | 1.14 (0.97, 1.33) | 0.106 | n/a |
| Total PACCs (per 1 PACC) | 1.22 (1.05, 1.42) | 0.011 | n/a |
| **Multivariable** |  |  |  |
| CAPRA-S (per 1 unit) | 1.08 (0.93, 1.27) | 0.322 | 0.075 |
| Total PACCs (per 1 PACC) | 1.17 (0.995, 1.38) | 0.057 |  |

Abbreviations: HR, hazard ratio; CI, confidence interval; LRT, likelihood ratio test; n/a, not applicable
\* The likelihood ratio test is based on the change in deviance when total PACCs is added to a model of CAPRA-S alone.

## Discussion

The major cause of death related to PCa is the development of therapy resistant metastatic disease. The possible mechanisms of therapy resistance in PCa have been broadly investigated with multiple candidates such as *SOX2* activation, *MYC* and *RAS* co-activation, and *ERG* gene rearrangements ^45–47^. However, the PACC state may represent an inclusive and unifying explanation for therapy resistance mechanisms that is under-recognized. This cell state is induced by the tumor microenvironment or therapeutic stress, can exist for an extended period of time, and can act as a CSC by undergoing depolyploidization and repopulating the tumor cell population when stress is relieved. In order to systematically study this phenomenon, one needs to go back to the fundamental approach to tumor pathogenesis: cell morphology.

PACCs have two defining characteristics: polyploidy and relatively large size. Polyploidy does not necessarily mean “multi-nucleation” and can be pronounced as a single large nucleus; however, multi-nucleated cells are often polyploid. Because of the increased genomic content, polyploid cells are physically larger than the neighboring tumor cells ^12^. The presence of PACCs has been shown to be associated with worse prognosis, higher tumor grade, poor differentiation, and advanced disease stage in various tumor types including PCa ^30,32,34,36,37,40^. There is also evidence in castration-resistant PCa that PACCs drive resistance to taxane-based chemotherapy^48^.

In this study, we investigated the presence of cells in the PACC state and their clinical importance in patients who underwent radical prostatectomy with curative intent to treat their presumed localized PCa. Our findings show that the number of PACCs and the number of cores positive for PACCs are statistically significant prognostic factors for metastasis-free survival, after adjusting for CAPRA-S, in a case-cohort of intermediate or high-risk men who underwent radical prostatectomy. In addition, despite the small number of men with complete data to evaluate time to mCRPC, the total number of PACCs was a statistically significant predictor of mCRPC in univariate analysis, and suggested a prognostic effect even after adjusting for CAPRA-S. To our knowledge, our study is the first to describe the adverse clinical implications of the presence and amount of PACCs in a stratified cohort of PCa patients based on the metastasis status. Assessing the prognostic value of PACCs for mCRPC by employing a larger cohort, prospective analyses of the predictive value of PACCs for adverse clinical outcome, and, ultimately, whole genome and RNA sequencing of the genetic material of these cells by using microdissection methods are of interest in understanding the biology of PACCs. It should be pointed out, however, that none of the PACCs-associated variables in **Table 2** was as strong as Gleason score, pathological stage or CAPRA-S.

One of the main challenges in this study was to accurately identify PACCs. We found the EpCAM stain helpful in visualizing the larger and atypical tumoral cells and increasing our ability to detect PACCs of various morphologies. It is clear that using a specific biomarker to highlight PACCs would be the ideal approach in studying these cells. However, there are currently no biomarkers for PACCs, either for monitoring *in vivo* or for isolation, and we believe this should be an area for future research.

Another challenge that might have affected the results of this study is tumor heterogeneity, which has long been known to be present in prostate cancer ^49^, given the fact that we employed TMAs to detect PACCs in our cohort. Although we applied a 3-4 fold sampling redundancy to reduce the margin of error, it is clear that the results of this study might have been affected by r under-representation of PACCs because of the heterogeneous nature of prostate cancer.

We did not determine the incremental improvement in the concordance index associated with adding PACCs to the model containing CAPRA-S because we aren’t aware of a validated approach to doing so for a case-cohort study design. However, it has been shown that if a variable is a statistically significant addition to a multivariable model it is mathematically equivalent to demonstrating a significant improvement in model performance, and that the test for significance of adding the variable has greater statistical power than a test of increase in concordance index. Since PACCs was statistically significant when added to a model with CAPRA-S it implies that adding the biomarker significantly improved model performance ^50,51^.

Because of their important role in disease resistance, we believe it is important to eliminate PACCs in treatment of aggressive PCa patients. However, there are no agents to specifically target these cells to date. Their unique biology and phenotype may create therapeutic opportunities as they may have unexpected vulnerabilities. This is a critical area of research in combination with molecular analysis of their genome. Although there is scarce knowledge about the biology of PACCs, we and others have shown their likely role in mediating disease resistance. We present additional evidence that they are significant prognostic factors for metastasis in patients with PCa who underwent radical prostatectomy with curative intent to treat their presumed localized PCa.

## Data Availability

All data produced in the present study are available upon reasonable request to the authors.

---

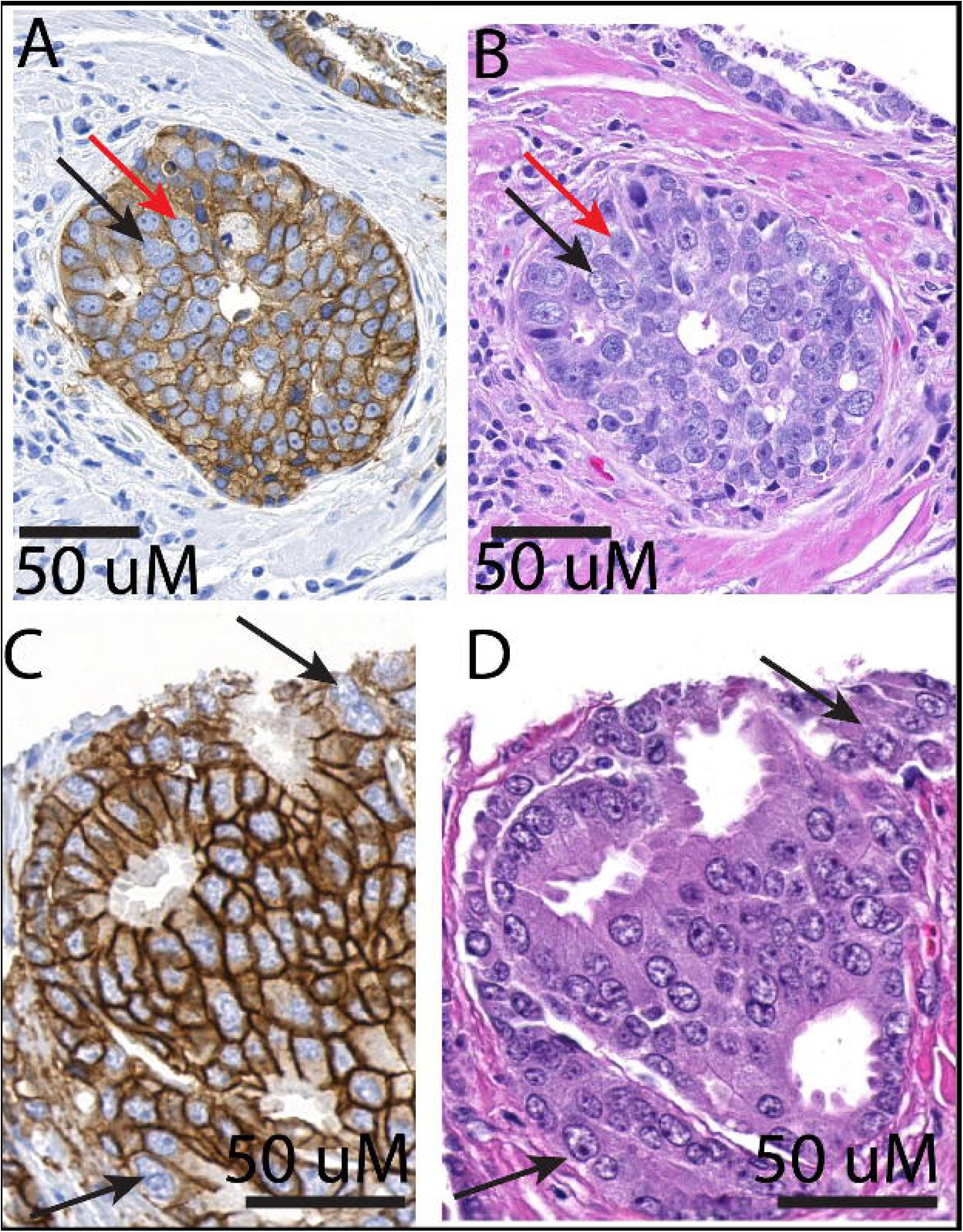

